# The health effects of 14 weeks of physical activity in a real-life setting for adults with intellectual disabilities

**DOI:** 10.1101/2022.05.17.22272069

**Authors:** Laurits Munk Højberg, Eva Wulff Helge, Jessica Pingel, Jacob Wienecke

## Abstract

**Background:** Individuals with intellectual disabilities (ID) have a reduced physical health compared to the general population, and one of the main contributors is inactivity.

**Aim:** To investigate how 14 weeks of physical activity (PA) in a real-life setting affects cardiovascular fitness, body composition and bone health of adults with ID.

**Methods:** Adults with ID were recruited into a PA-group or a control group (CON). The PA-group participated in 14 weeks of PA. Body composition, cardiovascular fitness and bone health were assessed before and after the intervention.

**Outcomes & results:** Cardiovascular fitness and body composition improved from pre to post within the PA-group: Heart rates (HR) during the last 30 seconds of two increments of a treadmill test, were reduced (3.2 km/h: -4.4 bpm, p<0.05; 4.8 km/h: -7.5 bpm, p<0.001) and fat mass was reduced (−1.02 kg, p<0.05). Between-group differences in favour of the PA-group, were observed in whole body bone mineral density (BMD) (0.024 g/cm^2^, p<0.05) and in BMD of the left femur neck (0.043 g/cm2, p<0.05).

**Conclusions & Implications:** Fourteen weeks of PA increase cardiovascular fitness, reduced fat mass and improved BMD in the weight-bearing skeleton in the PA-group. Increased and regular PA seems to be a promising tool to promote physical health in adults with ID.

**What this paper adds:** This paper underlines the importance of including physical activity in the everyday lives of individuals with intellectual disabilities. The health impact of physical activity performed outside controlled research laboratories needs further investigation, and therefore this paper sheds light on physical activity performed in a real-life setting. Our results indicate a positive impact of physical activity on the cardiovascular system and body composition, as well as bone health. The question of how of bone health of adults with intellectual disabilities responds to physical activity is largely understudied in intervention designs, and this paper includes state-of-the art investigation of development in bone mineral density after participation in varied physical activities. These results give promise and indicate a positive impact of the intervention on the bone health of the participants.

## 1. Background

Individuals with intellectual disabilities (ID) are subjected to increased mortality, in Denmark, individuals with ID die 14.5 years earlier than the general population (Flachs, Uldall, & Juel, 2014). The increased mortality is in part attributed to decreased physical health, high levels of inactivity and obesity which can lead to lifestyle diseases such as type II diabetes, cardiovascular disease and poor bone health (Anderson et al., 2013; Bartlo & Klein, 2011; Melville, Hamilton, Hankey, Miller, & Boyle, 2007; Robertson et al., 2000; Srikanth, Cassidy, Joiner, & Teeluckdharry, 2011; Zylstra, Porter, Shapiro, & Prater, 2008). Physical activity (PA) has been suggested as an effective way to improve the health in individuals with ID (Bartlo & Klein, 2011; Bouzas, Ayán, & Martínez-Lemos, 2018; Robertson et al., 2000). Previous investigations of individuals with ID have shown that the group respond to physical activity similarly to the general population. Evidence from controlled trials and systematic reviews show improvements in cardiovascular fitness following 80-135 minutes of aerobic training per week for 8-16 weeks (Boer et al., 2014; Cluphf, O’Connor, & Vanin, 2001; Dodd & Shields, 2005). The evidence regarding resistance training indicates that 8-24 weeks of resistance training increases muscle strength of persons with ID (Cowley et al., 2011; Obrusnikova, Firkin, Cavalier, & Suminski, 2021; Shields et al., 2013). Interventions with both aerobic and resistance training has shown that the participants experienced the benefits from both these modalities, within 10-20 weeks (Calders et al., 2011; Elmahgoub et al., 2011; Oviedo, Guerra-Balic, Baynard, & Javierre, 2014). PA for individuals with ID seem to induce reductions in body weight and fat mass (Elmahgoub et al., 2011; Son, Jeon, & Kim, 2016), and improved bone health (González-Agüero et al., 2012; Lizondo, Caplliure-Llopis, Escrivá, De La Rubia, & Barrios, 2019), but the results are equivocal within these areas, and more research is warranted. Improvements in the above-mentioned domains have a positive influence on general health. Improved cardiovascular fitness is associated with reduced risk of early death from cardiovascular and respiratory diseases (Steell et al., 2019), and increased muscle strength in the lower body can prevent falls and fractures (Benichou & Lord, 2016). The mentioned studies have generally used supervised exercise interventions, with researchers controlling the PA performed by the participants. The evidence from these studies are fundamental, but the translation into real-life, health-promoting exercise programs for the population with ID is difficult and demand further research (Bartlo & Klein, 2011; Bouzas et al., 2018). Hence, the aim of the present study was to investigate how 14 weeks of varied and adapted PA performed in a real-life setting, 5 days per week affects the health of adults with ID. To fulfil this aim, we collaborated with a local daily activity centre, where adults with ID is enrolled for a year, and participating in PA at the centre 5 days a week. Thus, an already feasible intervention could be investigated on a large number of participants. We hypothesised that a PA-group would improve their health from pre-to post-test compared with a control group. In addition, we hypothesized that a group of participants, who had already attended the activity centre at least one school year prior to the study (2^nd^-year participants), would have a better health profile at PRE-tests than the participants, who started their first year at the centre when entering the study (1^st^-year participants), due to impact of the PA that the 2^nd^-year participants had already performed. For the same reason, we expected to find the largest improvements in the selected health parameters in the 1^st^-year group over the course of the study.

## 2. Methods

### 2.1 Intervention and setting

The intervention consisted of varied forms of PA, performed in four periods of 3-4 weeks in duration, for a total of 14 weeks. Period 1: Dance, cycling, basic ballgames, outdoor fitness and athletics; Period 2: Adventure, motor training, football (soccer) and maritime outdoor activities. Period 3: Body theatre, swimming, floorball, l and E-sport. Period 4: Body theatre, racket sports, volleyball and fitness. The activities were performed at the activity centre; the Sports School for Adults with Developmental Disabilities (SSADD) in Copenhagen, Denmark. Adults with ID attending the SSADD, referred to as “participants”, were recruited into the PA-group. Each participant chose one of the above activities, in each period. Five days per week, they had 150 minutes of scheduled PA, in two blocks of 75 minutes with 45 minutes of break time in between. The SSADD has a capacity of ∼50 adults with different grades of ID, per school year. Each year approximately half of the participants continue at SSADD for an additional year (2^nd^ year participants), and ∼25 new participants start at the school (1^st^ year participants). The personnel at SSADD aims at improving the physical, social and personal competences of the individuals attending the institution, through the use of inclusive PA and sports, and the activities are adjusted to meet the capabilities of the individuals attending the SSADD.

### 2.2. Participants

Fifty-two adults with ID (26.7 ± 1.2 years) were recruited from the SSADD (PA-group). The PA-group was further divided into a 1^st^-year group and a 2^nd^-year group. A control group (CON-group) of 14 adults (29.1 ± 1.7 years) were recruited from other institutions and activity centres in Copenhagen. The institutions which the individuals in the CON-group attended did not focus on physical activity, but were recreationally physically active (Figure 1). This study was carried out with approval of The Danish Committee on Health and Research Ethics (H-19017828). All participants were thoroughly informed about the study (one-to-one) by a member of the research team, where they were made aware of their rights as participants, and were able to ask questions before giving their written consent to participate in the present study. Family members or guardians were present when needed to support the conversation. Being under legal guardianship was an exclusion criteria. The study followed with the Helsinki declaration. Since the project ran for two years, some participants were tested PRE and POST twice, with a year between the intervention periods. Between the two rounds of measurements, the subjects were allowed to change groups; for example, many participants the 1^st^-year group in the first round continued at SSADD, and were then included in the project in the 2^nd^-year group (Figure 1). Some participants who had initially given their consent to participate in the study chose to withdraw their consent, during the course of the study. These circumstances resulted in different numbers of participants both between the different tests and between the PRE and POST measurements. An overview of the number of measurements performed in the different tests can be found in Appendix 1. The grade of ID of the participants varied from mild to moderate and consisted of individuals with diagnoses such as Downs’ syndrome, cerebral palsy and other variants of ID, which the researchers did not have information on.

**Figure 1:**
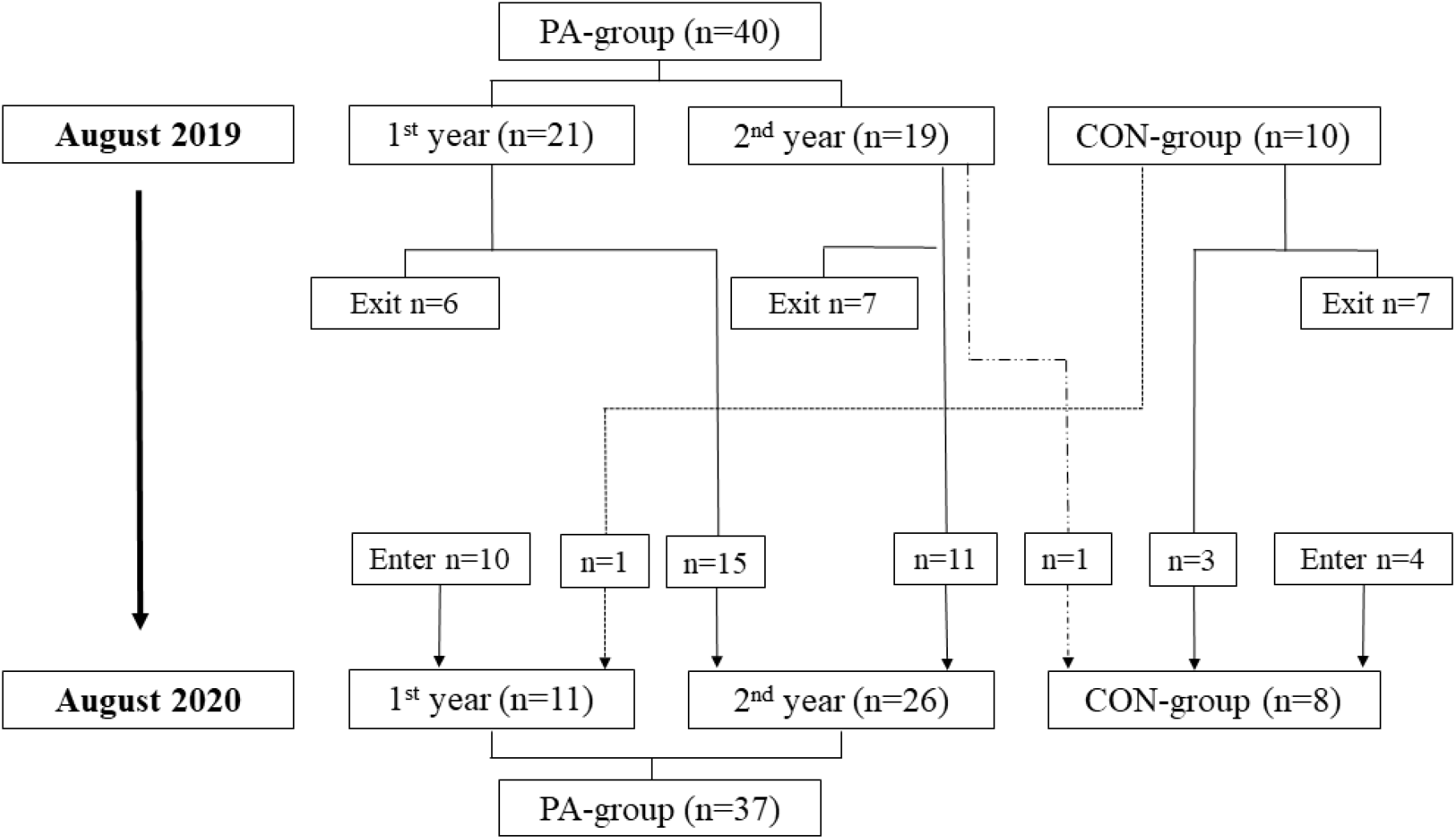
Flow chart of the movements between the groups of the participants between the two rounds of experiments, from August 2019 to August 2020.

### 2.3. Testing procedure

The individuals in the PA-group were recruited during their first week at SSADD over two intervention rounds, in 2019 and 2020. After recruitment, the participants in the PA-group underwent the tests described below over the course of two weeks. Participants in the CON-group were recruited during this period, and were tested after the PA-group. From PRE-to POST-tests, 14 weeks passed on average in both groups. All tests were carried out at the facilities of the Department of Nutrition Exercise and Sports, University of Copenhagen.

### 2.4. Assessment of physical activity

To measure the total amount of daily PA performed by the participants, accelerometry using the Axivity AX3 system were performed. Accelerometers (Axivity, Newcastle UK) were attached to the right thigh of the participants for periods of seven days, using medical tape and waterproof artificial skin (Opsite Flexifix ®). The accelerometers were checked daily in the weekdays to ensure correct positioning. If the accelerometer had detached it was reattached and recalibrated. Based on the threshold values of standard deviation of acceleration and the derived deviation (Skotte, Korshøj, Kristiansen, Hanisch, & Holtermann, 2014), the software ACTi4 (ACTI Corp., Copenhagen, Denmark) was used to discriminate the accelerometer data into different PA types (sitting, standing, walking, walking on stairs, running, cycling and total number of steps). This enabled the measurement of steps taken pr. hour and minutes of activity pr. week. The participants were asked to register their sleep schedules (hours slept) with the help of family or personnel at their home. Forty-three participants were divided between six different activities; football (n=9), motor training (n=5), fitness activities (n=9), outdoor activities (n=13) and cycling (n=7). Not all participants were able to wear the accelerometer for seven days. In these cases, the days they had completed, were included in the data.

### 2.5. Dual X-ray Absorptiometry (DXA) scans

The outcome measures of interest to the present study was areal bone mineral density (BMD, g/cm^2^), and body composition measures (e.g. fat mass (kg) and fat free mass (kg)). The DXA-protocol consisted of a whole body scan (Albanese, Diessel, & Genant, 2003) and scans of the lumbar vertebrae (L1-L4) and bilateral proximal femur, in that order (Lunar iDXA, GE Healthcare). DXA-scans of the lumbar and femur regions are routinely used as clinical tools for diagnosing osteoporosis (Blake & Fogelman, 2007).. The participants’ weight and height were recorded prior to the scans (Soehnle Professional Scale, Soehnle, Germany). The participants were asked to remove all metal objects (jewellery, buttons etc.) from their body and clothing prior to the scans. The participants were asked to arrive to the tests in a fasting state. Not all participants were able to meet this request, so their fasting state was recorded at the PRE-tests (fasting/non-fasting). The participants were asked to arrive to the POST-test in the same fasting-state as at the PRE-test. The participants were placed in the middle of the scanning bed with their arms held to the sides of the body, their hands in neutral position and their palms on the side of their thighs. With the knees bent in a 90 degrees angle, a foam block was placed under the participant’s calves for the lumbar scans to ensure that the lower back was placed correctly on the scanning bed. During the femur scans, the participants were asked to rotate their legs inwards from the hips, while their feet were strapped to a plastic trapezoid. This was done to ensure the right position of the femur neck, and to promote reproducibility of the measurements.

### 2.6. Oxygen uptake protocol and heart rate

The submaximal oxygen uptake (VO2)-protocol of the present study was inspired by study by Ebbeling and colleagues (Ebbeling, Ward, Puleo, Widrick, & Rippe, 1991), where a submaximal treadmill test for estimation of VO_2_max was developed. The test consisted of a 4 minute warm up with 0% incline at two different speeds, two minutes at 3.2 km/h followed by two minutes at 4.8 km/h. The warm-up was followed by 1 minute at standing rest. After the rest the participants performed the test, a bout of 8 minutes with two intensities. First, the participants walked at 3.2 km/h and 5% incline for 4 minutes, immediately followed by 4 minutes at 4.8 km/h and 5% incline. Online breath-by breath analysis was performed during the exercise protocol using a metabolic cart (Masterscreen CPX, CareFusion). Participants wore a mask with ∼70 ml dead space, which was directly connected to the gas analyser and flow sensor. The system was calibrated before each trial. The heart rates (HR) were sampled using POLAR H3 heart rate sensors. An experimenter noted the heart rates of the participants every 10 seconds during the exercise protocol. The area under the curve (AUC) of the VO_2_-curves during intensity transitions is an estimate of oxygen uptake kinetics. The first 20 seconds of the transitions were omitted from the analyses to exclude phase 1 kinetics and investigate phase 2 kinetics. AUC were then calculated for the subsequent 100s. An increase in AUC of the VO_2_-curve during similar intensity transitions and a fixed time frame, is a measure of increased total oxygen consumption.

### 2.7. Data analysis and statistics

Data was analysed using R statistics (*R statistics 4*.*0*.*0*, 2020). Linear mixed models (LLM’s) were used to analyse changes within and between groups (“lme4” R-package (Bates, Mächler, Bolker, & Walker, 2015)). In the first LLM, the group variable had two levels; PA and CON. In the second LLM, the group variable had three levels; 1^st^-year group, 2^nd^-year group and CON. The LLM’s included the age, gender and diagnosis of the participants as well as the test round (i.e. 2019 and 2020), as fixed effects, and the participant ID’s as a random effect. LLM’s were chosen, for their ability to handle missing data. Post hoc analysis of the models were used to investigate the changes from PRE-to POST-tests between and within the groups (“multcomp” R-package (Hothorn, Bretz, & Westfall, 2008)). The p-values were corrected for multiple comparisons with the single step method. The critical value for statistical significance were assumed at an alpha level p≤0.05. Unless otherwise stated, data is presented as mean ± standard error (SE). Percent changes within groups are calculated as the change from PRE-to POST-test, and percentwise differences between groups are calculated as the change score between the PA groups and the CON group, relative to the CON-group PRE-test value.

## 3. Results

### 3.1 Demographic data

The demographic data collected at the PRE-measurements is presented in Table 1. Some participants were tested in both 2019 and 2020. They have contributed to the demographic data once, in 2019. An overview of the total number of data points included in the measurements, can be found in Appendix 1.

**Table 1:** Demographic data of participants from their first measurement

| Variables | PA | 1st | 2nd | CON |
| --- | --- | --- | --- | --- |
| Demographic data |  |  |  |  |
| Number of participants | 52 | 32 | 20 | 14 |
| Gender (f/m) | 28/24 | 17/15 | 11/9 | 5/9 |
| Age (years) | 26.7 ± 1.2 | 26.7 ± 1.5 | 26.7 ± 1.4 | 29.1 ± 1.7 |
| Height (cm) | 163.3 ± 1.8 | 162.3 ± 2.2 | 165.1 ± 3.0 | 163.3 ± 2.4 |
| Developmental disorder (DS/CP/UD) | 15/6/31 | 12/3/19 | 5/3/12 | 4/1/9 |

The number of participants in the study groups. The participants' age was recorded at PRE-
tests. Abbreviations: f: female, m: male, cm: centimetres, DS: Downs' Syndrome, CP: Cerebral
Palsy, UD: Undefin

### 3.2 Daily physical activity

During one week, the participants in the PA-group on average took more steps/hour (p<0.001) while at SSADD (1453 ± 60 steps/h) compared to leisure hours, both on weekdays (738 ± 67 steps/h) and in the weekends (564 ± 58 steps/h) (Figure 2). The PA-group performed on average 407 minutes of PA/week during the total of 1500 minutes they spent at the school. This included activities other than standing, sitting or lying down. As mentioned, they had 150 minutes of scheduled PA/day (equivalent of 750 minutes/week).

**Figure 2:**
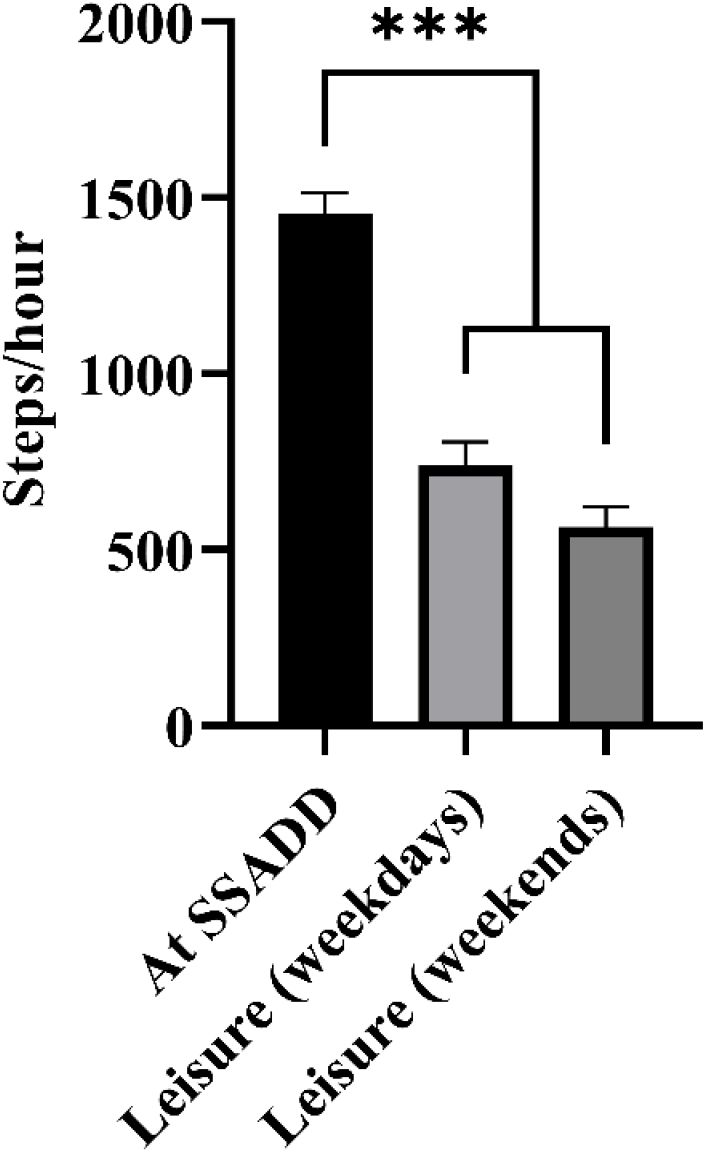
The amount of steps taken pr. hour while the participants were at SSADD, during leisure on weekdays after the school day ends and during weekends. Measurements were made over a 7 day period. Error bars: Standard error of the mean. Significance codes: p<0.001: “***” 0.001>p<0.01: “**”, 0.01>p<0.05: “*”, 0.05>p<0.1: “.”

### 3.3 PRE-test measurements

Raw PRE-test values are presented in Table 2. The comparison between the 1^st^-year group and the 2^nd^-year group showed a higher total body weight (4.7%, p<0.05) and a higher fat mass (11%, p<0.001) in the 1^st^-year group. In addition, the 1^st^-year group had a higher visceral (16.2%, p<0.001) and gynoid fat mass (8.9%, p<0.05) than the 2^nd^-year group. The 1^st^-year-group had a higher heart rate (HR) compared to the 2^nd^-year group during the last 30s at 3.2 km/h (5.0%, p>0.05) (Figure 3). The PA-group had lower BMD compared to the CON-group in L1-L4 (−6.8%, p>0.05). The T-score distribution of the right femur neck of the PA-group was: T ≤ -2.5 (osteoporosis): 0%, -2.5 > T ≤ -1 (osteopenia): 23.5%, -1 > T < 0: 29.4% and T ≥ 0: 43.1 %. The CON-group distribution was: T ≤ -2.5 (osteoporosis): 0%, -2.5 > T ≤ -1 (osteopenia): 28.6%, -1 > T < 0: 35.7% and T ≥ 0: 35.7%. As no differences were observed between the right and left sides at PRE-tests, only data from the right side is presented.

**Table 2:**
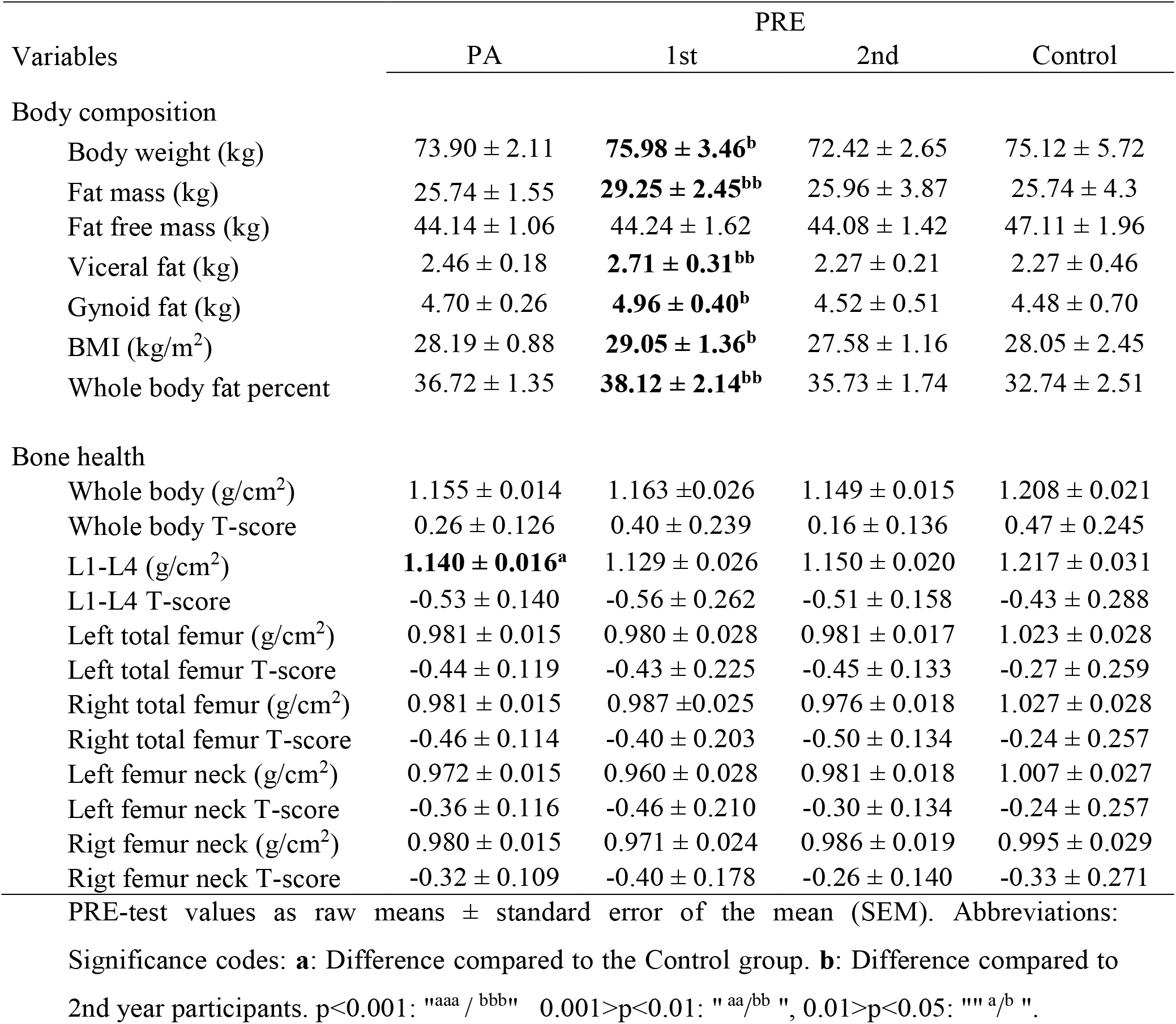
Body composition and bone health measures at PRE-tests

| Variables | PA | PRE |  |  |
| --- | --- | --- | --- | --- |
|  |  | 1st | 2nd | Control |
| Body composition |  |  |  |  |
| Body weight (kg) | 73.90 ± 2.11 | **75.98 ± 3.46<sup>b</sup>** | 72.42 ± 2.65 | 75.12 ± 5.72 |
| Fat mass (kg) | 25.74 ± 1.55 | **29.25 ± 2.45<sup>bb</sup>** | 25.96 ± 3.87 | 25.74 ± 4.3 |
| Fat free mass (kg) | 44.14 ± 1.06 | 44.24 ± 1.62 | 44.08 ± 1.42 | 47.11 ± 1.96 |
| Visceral fat (kg) | 2.46 ± 0.18 | **2.71 ± 0.31<sup>bb</sup>** | 2.27 ± 0.21 | 2.27 ± 0.46 |
| Gynoid fat (kg) | 4.70 ± 0.26 | **4.96 ± 0.40<sup>b</sup>** | 4.52 ± 0.51 | 4.48 ± 0.70 |
| BMI (kg/m<sup>2</sup>) | 28.19 ± 0.88 | **29.05 ± 1.36<sup>b</sup>** | 27.58 ± 1.16 | 28.05 ± 2.45 |
| Whole body fat percent | 36.72 ± 1.35 | **38.12 ± 2.14<sup>bb</sup>** | 35.73 ± 1.74 | 32.74 ± 2.51 |
| Bone health |  |  |  |  |
| Whole body (g/cm<sup>2</sup>) | 1.155 ± 0.014 | 1.163 ± 0.026 | 1.149 ± 0.015 | 1.208 ± 0.021 |
| Whole body T-score | 0.26 ± 0.126 | 0.40 ± 0.239 | 0.16 ± 0.136 | 0.47 ± 0.245 |
| L1-L4 (g/cm<sup>2</sup>) | **1.140 ± 0.016<sup>a</sup>** | 1.129 ± 0.026 | 1.150 ± 0.020 | 1.217 ± 0.031 |
| L1-L4 T-score | -0.53 ± 0.140 | -0.56 ± 0.262 | -0.51 ± 0.158 | -0.43 ± 0.288 |
| Left total femur (g/cm<sup>2</sup>) | 0.981 ± 0.015 | 0.980 ± 0.028 | 0.981 ± 0.017 | 1.023 ± 0.028 |
| Left total femur T-score | -0.44 ± 0.119 | -0.43 ± 0.225 | -0.45 ± 0.133 | -0.27 ± 0.259 |
| Right total femur (g/cm<sup>2</sup>) | 0.981 ± 0.015 | 0.987 ± 0.025 | 0.976 ± 0.018 | 1.027 ± 0.028 |
| Right total femur T-score | -0.46 ± 0.114 | -0.40 ± 0.203 | -0.50 ± 0.134 | -0.24 ± 0.257 |
| Left femur neck (g/cm<sup>2</sup>) | 0.972 ± 0.015 | 0.960 ± 0.028 | 0.981 ± 0.018 | 1.007 ± 0.027 |
| Left femur neck T-score | -0.36 ± 0.116 | -0.46 ± 0.210 | -0.30 ± 0.134 | -0.24 ± 0.257 |
| Rigt femur neck (g/cm<sup>2</sup>) | 0.980 ± 0.015 | 0.971 ± 0.024 | 0.986 ± 0.019 | 0.995 ± 0.029 |
| Rigt femur neck T-score | -0.32 ± 0.109 | -0.40 ± 0.178 | -0.26 ± 0.140 | -0.33 ± 0.271 |

PRE-test values as raw means ± standard error of the mean (SEM). Abbreviations:
Significance codes: **a**: Difference compared to the Control group. **b**: Difference compared to
2nd year participants. p<0.001: "aaa / bbb" 0.001>p<0.01: "aa/bb ", 0.01>p<0.05: "" a/b ".

**Figure 3:**
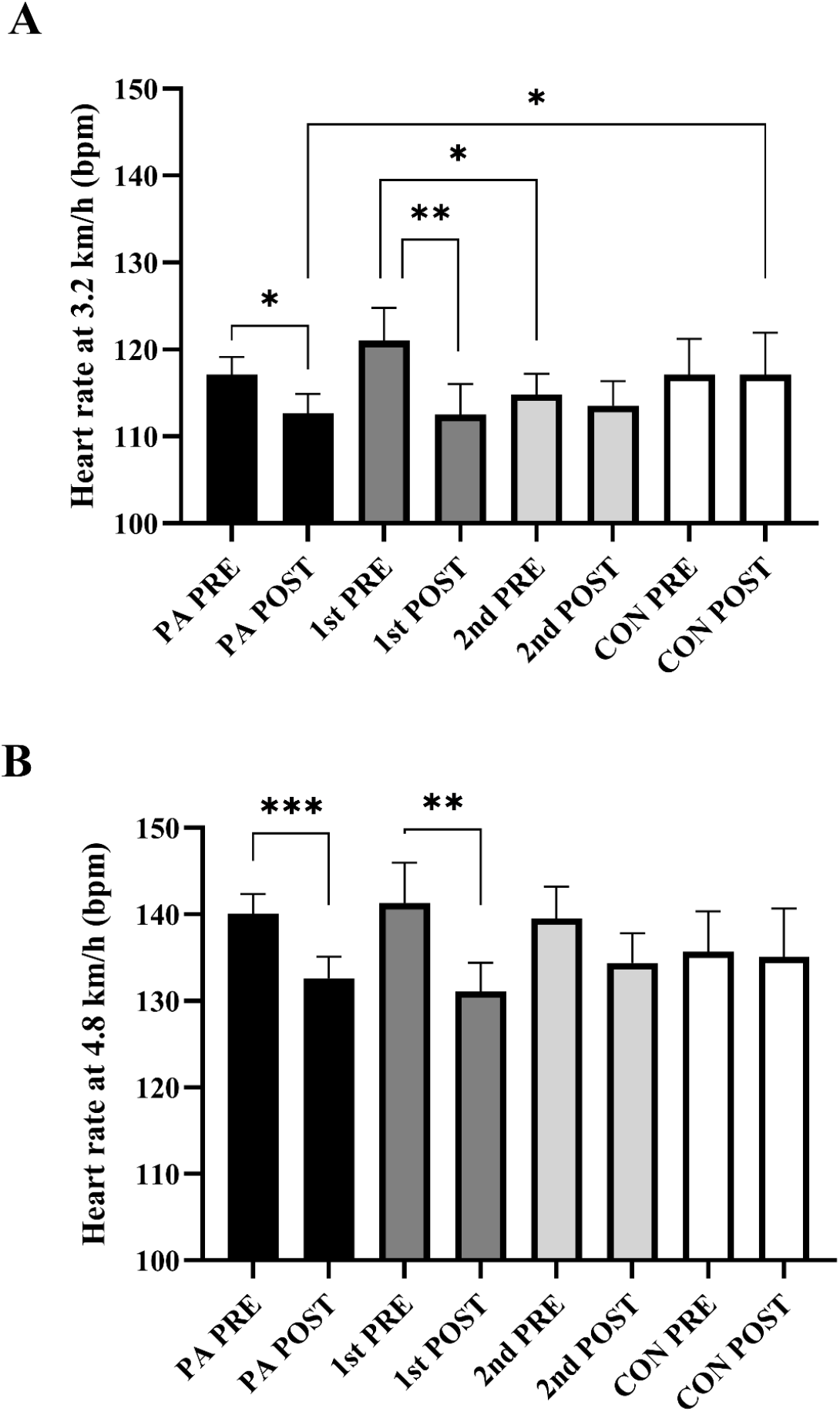
The average heart rates during the last 30 s. at 3.2 km/h (A) and 4.8 km/h (B) for the PA-, 1^st^-year, 2^nd^-year and CON-group. Significance codes: p<0.001: “***” 0.001>p<0.01: “**”, 0.01>p<0.05: “*”, 0.05>p<0.1: “.

### 3.4 Pre-to posttest changes

#### 4.4.1 Body composition

Table 3 presents the model estimated Δ values (POST – PRE) within the groups, and the differences in change scores between the groups (e.g. PA-group Δ value – CON-group Δ value). Analyses of body composition outcomes form the whole body DXA scans revealed that the total PA group lost -4.0% fat mass (p<0.05), with the 1^st^-year group lost -5.8% fat mass (p<0.05). For the 1^st^-year group, parts of the fat loss were located at the visceral and gynoid regions (−7.4% and -7.1%, p<0.05, respectively). However, no between group differences were observed (p>0.05).

**Table 3:**
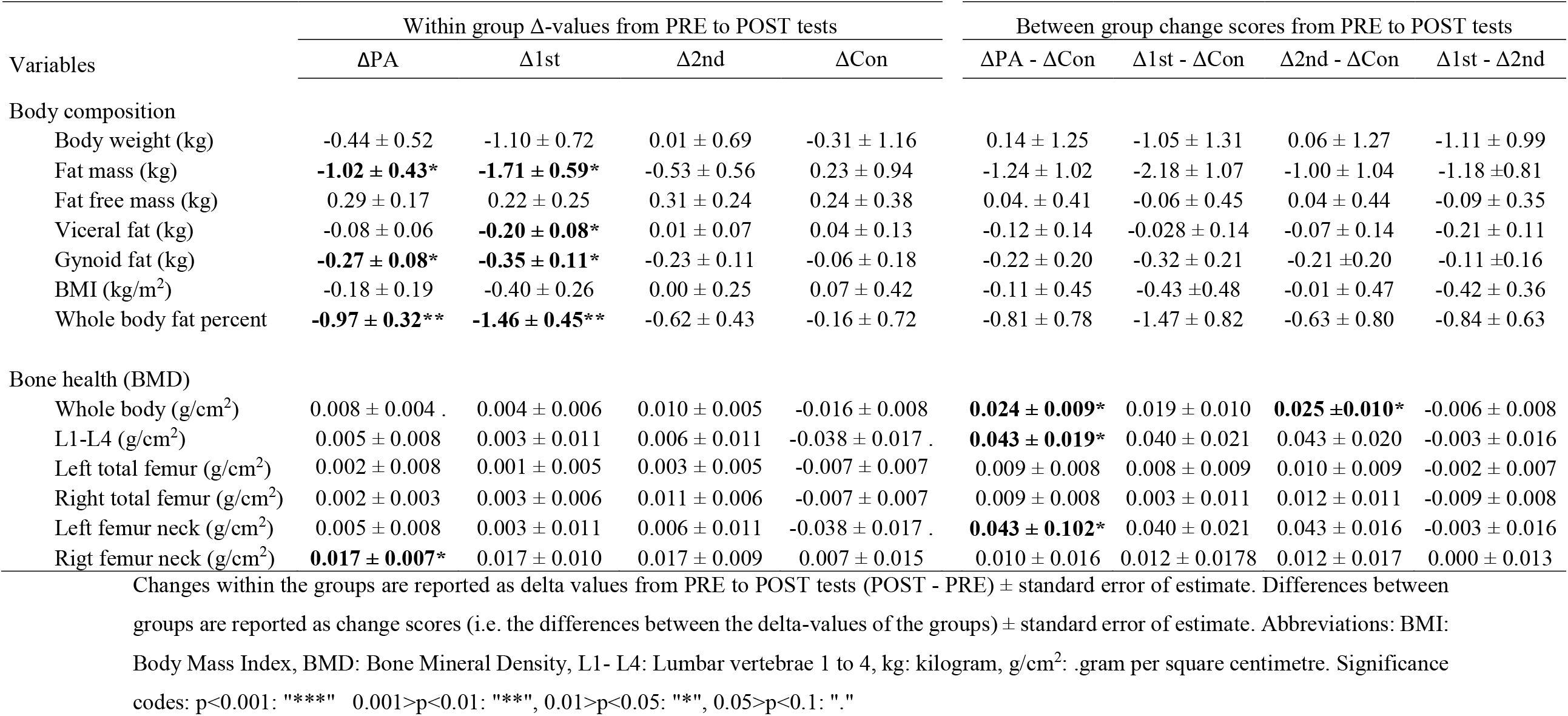
Changes within and between groups over time in body composition and bone health.

#### 3.4.2 Bone mineral density

A within-group change was observed in the PA-group, thus the BMD of the right femur neck increased (p<0.05) (Table 3). Tendencies indicating reduced BMD within the CON-group over the course of the study was evident as borderline significant reductions of the L1 and left femur neck (0.5< p ≤0.1). Differences in change scores for whole body BMD (2.0%, p<0.05) and BMD of the left femur neck (4.3%, p<0.05) between the PA- and CON-group, in favor of the PA-group, were observed.

#### 3.4.3 Cardiovascular fitness

Within-group changes was observed in the PA-group as a reduction in HR during the last 30s at both 3.2 km/h (−3.8%, p<0.05) and 4.8 km/h (−5.4%, p>0.001) (Figure 3). This decrease was more pronounced within the 1^st^-year group (3.2 km/h: -7.0%, p<0.01. 4.8 km/h: 7.2% p>0.001). However, no between group differences were observed in the HR measurements, and VO_2_-kinetics during the transitions of the treadmill test did not change (p>0.05).

### 3.5 Post-test differences

Between-group differences at POST-tests were analysed to investigate if the differences observed at PRE-tests were still present (data not shown). The differences in body composition observed at the PRE-tests between the 1^st^- and 2^nd^-year group, and the difference in BMD of L1-L4 between the PA-group and the CON-group (Table 2), were not observed at the POST-tests. The only statistically significant finding from this analysis was a significant 8.9 % lower HR at 3.2 km/h in the PA-compared to the CON-group (Figure 2).

## 4. Discussion

The results of the present study indicates that adults with ID by participation in PA in a real-life setting for 14 weeks can improve their body composition, bone health and cardiovascular fitness. The amount of PA performed by the PA-group is above the weekly 300 minutes of PA, recommended for adults by the World Health Organization (Bull et al., 2020).

### 4.1 Body composition

The PA-group lost 1.02 kg of fat mass and the 1^st^-year group lost 1.7 kg of fat mass, with no changes observed in body weight. The change in the total PA-group seems to be driven by the 1^st^-year group, as no changes were found for the 2^nd^-year group. The results of previous studies on the effect of physical activity on body composition for adults with ID are equivocal. Some studies on individuals with ID have reported reductions in fat mass of similar magnitude as the present study (Elmahgoub et al., 2011; Ordoñez, Rosety, & Rosety-Rodriguez, 2006; Wu et al., 2017) while other studies did not (Calders et al., 2011; Oviedo et al., 2014). In a meta-analysis of 6 RCT’s assessing the effects of PA interventions on body composition in young adults with ID, Harris and colleagues reported no overall effect of physical activity interventions on body composition (Harris, Hankey, Murray, & Melville, 2015). The studies included in the meta-analysis used interventions of 80-195 minutes of activity/week (Donnelly et al., 2009; Harris et al., 2015). As Harris and colleagues suggested, this might be too little to elicit changes in body composition. In the present study, the participants in the PA-group performed ∼400 minutes of PA/week, of variable intensity. Thus, the training load imposed on our participants, should be enough to elicit the adaptations in body composition observed in the present study. This is supported by the findings from a systematic review on the effects of exercise on fat mass in the general population, where ∼10 weeks of either moderate-or high-intensity exercise, induced ∼2 kg fat mass reduction (Wewege, van den Berg, Ward, & Keech, 2017). Food intake is another important variable when studying changes in body composition (Clifton, Condo, & Keogh, 2014; Miller et al., 2013). Studies on individuals with ID have successfully improved body composition through multi-component interventions with exercise, diet and motivation (Martínez-Zaragoza, Campillo-Martínez, & Ato-García, 2016). The diet of the participants were not monitored in the present study. Thus, the results indicate that it is possible to reduce fat mass in this population, solely with increased PA without a diet intervention. Significant parts of the fat mass reduction seen in the present study could be located to the gynoid and visceral areas (Table 3). The visceral fat mass reduction for the 1^st^-year group is of significance in a health perspective, as the amount of visceral fat is a risk factor in relation to life style diseases such as type II diabetes mellitus and cardiovascular diseases (Finelli, Sommella, Gioia, La, & Tarantino, 2013; Kuk et al., 2006; Lee et al., 2018). Thus, our results show a positive effect of 14 weeks of PA performed at SSADD on fat mass.

### 4.2 Bone health

All together, the between group differences in BMD observed in whole body, L1-L4 and the left femur neck indicate that the PA-group benefitted from 14 weeks of PA, compared to the CON-group (Table 3). The differences between the groups seem to be driven by a decrease in BMD in the CON-group, while the PA-group maintained their BMD in these regions. Another indication of this, is the higher BMD of L1 -L4 of the CON-group compared to the PA-group at PRE-tests (Table 2), which were not present at POST-tests (data not shown).

To our knowledge, the present study is one of the first to investigate the bone health in individuals with ID after a PA intervention, and the first to do so in adults. A previous study on 7-10 year old boys with ID found an increase in femoral neck BMD after 6 months of exercise (165 min/week) (Hemayattalab, 2010). Another study observed an increase in bone mineral content of the femur in a group of subjects with DS between 10 and 19 years, after 21 weeks of exercise (50-60 min/week) (González-Agüero et al., 2012). The results of the present study is thus in line with the previous research, showing that PA has an osteogenic effect on individuals with ID. In the present study, the osteogenic effect of PA is evident as an increase in right femur neck BMD in the PA-group, and as a counteraction of decrease in lumbar BMD in the PA-group relative to the CON-group. There are differences between the present study and earlier research regarding bone adaptations to exercise. First, it is common in this line of research to include activities aimed at inducing an osteogenic response; activities which imposes strain and impact to the bones (Ehrlich & Lanyon, 2002; Hinton, Nigh, & Thyfault, 2015; Kato et al., 2006). The study by Gonzalez-Agüero and colleagues used plyometric training and the study by Hemayatta’s lab used running, hopping and skipping exercises. In the present study, we did not monitor what type of activities were performed, and the accelerometers used were not able to distinguish between activities at this level. Additionally, the present intervention had a duration of 14 weeks, which is a short time to observe substantial adaptations in bone structure as a result of training (Ahola, Korpelainen, Vainionpää, Leppäluoto, & Jämsä, 2009; Harding & Beck, 2017). Finally, previous studies investigated children and adolescents with ID, while this study concerned adults. With regards to the clinical relevance of the results, the increase of 0.017 g/cm^2^ of the right femur neck is within the range seen in systematic reviews of exercise studies on adults from the general population (Bolam, Van Uffelen, & Taaffe, 2013; James & Carroll, 2010), and it may be of significance to the fracture risk of the subjects (Kemmler, Bebenek, Kohl, & von Stengel, 2015). At PRE-tests, the bone health of the participants in the present study was in agreement with previous research, regarding the BMD of the whole body (Geijer, Stanish, Draheim, & Dengel, 2014) and of the lumbar vertebrae (Angelopoulou, Souftas, Sakadamis, & Mandroukas, 1999). Our study population had higher T-scores and healthier distribution of T-scores of the right femur neck, than what was observed in earlier cross-sectional studies on similar populations (Chen, Chen, Lin, & Chen, 2015; Zylstra et al., 2008). The lowest average T-score of the right femur neck in the present study was seen in the 1^st^-year group (−0.40), which was higher than the value reported by Zylstra and colleagues (−1.51) (Zylstra et al., 2008). Zylstra and colleagues reported an osteoporosis prevalence (T ≤ -2.5) in the femur region of 22% and an osteopenia prevalence (−2.5 > T ≤ -1) of 33% in 36 subjects with ID aged 19-29 years. The prevalence distribution of all participants in the present study was combined for comparison with the data reported by Zylstra and colleagues. We observed an osteoporosis prevalence of 0% and an osteopenia prevalence of 25.4 %. Zylstra and colleauges did not report which model of DXA-scanner they used in their study, which could influence the results. In summary, adults with ID is a population with low T-scores of the femur and lumbar regions, but the population in the present study are not in as high risk of developing osteoporosis, as earlier studies have reported (Jasien, Daimon, Maudsley, Shapiro, & Martin, 2012; Zylstra et al., 2008).

### 4.3 Cardiovascular fitness

The PA-group reduced their heart rate during the last 30 seconds on both speeds during the treadmill-test (Figure 3), but we did not find any changes in the amount of O_2_ consumed per kg body weight during the submaximal exercise transitions on the treadmill test. The reductions in HR point towards an improved cardiovascular function; the subjects’ hearts were able to meet the energy demand of the working muscles during the walking test with less beats per minute, possibly due to an increase in stroke volume (Hartley et al., 1969; Wilmore et al., 2001). The increase in cardiovascular fitness observed in the present study is in line with what was reported by earlier studies with similar target groups and intervention durations. Barwick and colleagues reported a decrease in HR in a after a 10 week functional training program (Barwick et al., 2012) and studies employing combined training programs observed increases in peak O_2_ uptake velocity (VO_2_peak) post intervention (Calders et al., 2011; Oviedo et al., 2014). The set-up of the present study complicates the comparison with the mentioned traditional training studies, especially with regards to the physical load imposed on the subjects. In the present study, we investigated the amount of PA performed by the PA-group while at SSADD and in their leisure (Figure 2), while traditional training studies delivered and monitored the PA conducted by the subjects. The knowledge obtained from these earlier studies about the effect of PA on cardiovascular fitness, together with the results of the present study, suggests a link between the PA performed at SSADD and the cardiovascular adaptations observed. Measures of cardiac function such as reduced resting HR and improved HR recovery reduces all-cause mortality from cardiovascular diseases in the general population (Adabag et al., 2008; Reil et al., 2011). We did not obtain these measures in the present study, but a reduced HR at the same absolute intensities could point towards an improved health-enhancing cardiac function.

### 4.4 Limitations

The quasi-randomized design of the present study poses a selection bias. Individuals who choose to attend SSADD could have an overall higher interest in PA, than the general population with ID. We tried to meet this challenge by including individuals in our control group, who were recreationally physically active in their leisure. The researchers observed the PA occasionally, but did not supervise the PA performed by the participants in the PA-group. Instead, this study focused on measuring the effects of an already existing PA option for the target group. Other studies have employed a similar approach, with regards to investigating already existing and well established offers and programs with PA for individuals with ID, and have found health-enhancing effects as well (Barak & Gan, 2019; Messiah et al., 2019; Rubenstein et al., 2020). This approach is valuable, when investigating the effects of existing solutions aimed upon the societal problem of the poor health status of individuals with ID. It would have been preferable to conduct accelerometry in the CON-group too, but this was not possible due to logistical challenges.

## 5. Conclusion

This study on the effects of 14 weeks of PA and sports participation for adults with ID observed positive health effects in the domains of body composition, bone health and cardiovascular fitness. The PA performed at SSADD seems to be most efficient in improving the health the of 1^st^-year group. For participants who have spent one or more years at the school (2^nd^-year group), the PA performed is likely to maintain their health in the parameters investigated in the present study. A continuously progressive increase in the intensity of the PA performed at SSADD could be a way to ensure continued health benefits for the participants in the 2^nd^ year-group. Improvements in the health parameters investigated in the present study could reduce the groups’ risk of developing lifestyle diseases.

## Data Availability

All data produced in the present study are available upon reasonable request to the authors

## Acknowledgements

The foundation of the project was the collaboration between the Department of Nutrition, Exercise and Sports, University of Copenhagen and the Sports School for Adults with Developmental Disabilities. Without the teamwork between the two institutions, this study would not have been possible. The data collection in this study, relied on the great help of Jens Jung Nielsen (academic staff), Rosita Olvhøj (scientific assistant), Kathrine Elsig Jensen (student) and Jonas Rud Bjørndal (PhD-student). Thanks to Peter Møller Christensen (Team Denmark), for guidance in the investigation of VO_2_ kinetics.

## Authors’ contributions

JW and LMH conceived the study. EWH supervised all work related to the DXA-scans. All authors contributed to the conception of the manuscript and approved the study before publication.

## Conflicts of interest disclosure

None of the authors have any financial or non-financial conflicts of interests, in relation to this study.

## Appendix 1

Participants, characteristics and distribution on the different outcomes.

| Tests | Numbers of measurements |  |  |  |  |  |  |  |
| --- | --- | --- | --- | --- | --- | --- | --- | --- |
|  | PA |  | 1st |  | 2nd |  | Control |  |
|  | PRE | POST | PRE | POST | PRE | POST | PRE | POST |
| DXA scans | 76 | 73 | 31 | 30 | 45 | 43 | 18 | 15 |
| HR (3.2 km/t) | 70 | 72 | 26 | 30 | 44 | 42 | 17 | 13 |
| HR (4.8 km/t) | 64 | 62 | 23 | 22 | 41 | 40 | 15 | 12 |
| Accelerometry | 43 | - | 21 | - | 22 | - | - | - |

| | Age (years ± SD) | | | | | | | |
| --- | --- | --- | --- | --- | --- | --- | --- | --- |
|  | PA |  | 1st |  | 2nd |  | Control |  |
|  | PRE | POST | PRE | POST | PRE | POST | PRE | POST |
| DXA scans | 26.7±8.3 | 26.8±8.0 | 26.8±9.0 | 27.0±9.2 | 26.7±8.0 | 26.4±7.1 | 28.9±5.8 | 29.7±6.2 |
| HR (3.2 km/t) | 26.7±7.8 | 26.5±8.0 | 26.7±7.6 | 27.1±9.1 | 26.7±8.0 | 26.0±7.1 | 28.9±6.0 | 28.7±4.7 |
| HR (4.8 km/t) | 27.1±8.1 | 26.5±7.5 | 27.3±7.9 | 27.3 ±8.1 | 26.9±8.3 | 26.1±7.2 | 29.7±5.9 | 28.1±4.5 |

|  | Gender (f/m) |  |  |  |  |  |  |  |
| --- | --- | --- | --- | --- | --- | --- | --- | --- |
|  | PA |  | 1st |  | 2nd |  | Control |  |
|  | PRE | POST | PRE | POST | PRE | POST | PRE | POST |
| DXA scans | 42/34 | 40/33 | 16/15 | 15/15 | 26/19 | 25/18 | 6/12 | 6/9 |
| HR (3.2 km/h) | 38/32 | 38/34 | 13/13 | 15/15 | 25/19 | 23/19 | 5/12 | 4/9 |
| HR (4.8 km/h) | 34/30 | 31/31 | 11/12 | 9/13 | 23/18 | 22/18 | 3/12 | 4/8 |

An overview of how the Abbreviations: DXA: Dual X-ray Absorptiometry HR: heart rate, km/h: kilometres per hour SD: standard deviation, f: female, m: male.

## Notes

**Ethics approval statement** This study was carried out with approval of The Danish Committee on Health and Research Ethics (H-19017828). All participants gave informed written consent and the study followed the Helsinki declaration. All participants were thoroughly informed about the study (one-to-one) by a member of the research team, where they were made aware of their rights as participants, and were able to ask questions before giving their written consent to participate in the present study. Family members or guardians were present when needed to support the conversation.

### Competing Interest Statement

The authors have declared no competing interest.

### Clinical Trial

NCT05336487

### Funding Statement

This research project was funded by the NovoNordisk Foundation and the Elsass Foundation.

### Author Declarations

This study was carried out with approval of The Danish Committee on Health and Research Ethics (case number: H-19017828)

